# An unbiased comparison of 14 epigenetic clocks in relation to 10-year onset of 174 disease outcomes in 18,859 individuals

**DOI:** 10.1101/2025.07.14.25331494

**Authors:** Christos Mavrommatis, Daniel W Belsky, Kejun Ying, Mahdi Moqri, Archie Campbell, Anne Richmond, Vadim N Gladyshev, Tamir Chandra, Daniel L McCartney, Riccardo E Marioni

## Abstract

Epigenetic Clocks have been trained to predict chronological age, healthspan and lifespan. Such clocks are often analysed in relation to disease outcomes – typically using small datasets and a limited number of clocks. Here, we present the first large-scale (n=18,849), unbiased comparison of 14 widely used clocks as predictors of 174 incident disease outcomes and all-cause mortality. Second-generation clocks significantly outperformed first-generation clocks, which have limited applications in disease settings. Of the 176 Bonferroni significant (P<0.05/174) associations, there were 27 diseases (including primary lung cancer and diabetes) where the hazard ratio for the clock exceeded the clock’s association with all-cause mortality. Furthermore, there were 35 instances where adding a clock to a null classification model with traditional risk factors increased the classification accuracy by >1% with an AUC_full_ > 0.80. Second-generation epigenetic clocks show promise for disease risk prediction, particularly in relation to respiratory and liver-based conditions.

## Main

We all age at the same chronological rate; everyone gets a year older every 12 months. But we experience the passage of time differently. Some of us maintain our health into our later years, while others develop chronic disease and disability by midlife. However, defining the biological differences that underlie this heterogeneity is complex. There is no gold standard measurement of biological aging [1]. However, there are now a number of candidate biomarkers of aging derived from integrating high-dimensional molecular data with machine-learning methods that have accumulated substantial validation evidence [2]. Among these, the best studied are a family of DNA methylation (DNAm) algorithms known as epigenetic clocks [3].

The first generation of epigenetic clocks were developed by comparing DNA methylation data between older and younger people to develop algorithms predictive of chronological age (e.g., the Horvath and Hannum clocks [4], [5]). A second generation of epigenetic clocks were subsequently developed by modelling differences between individuals in mortality risk and phenotypes that track general health [6], [7]. Finally, a third generation were developed by modelling differences between individuals of the same chronological age in their rate of multi-organ system deterioration [8]. The later generation clocks are therefore trained to predict complex outcomes, compared to chronological age. They often include two-step processes whereby DNAm proxies of health-related biomarkers (e.g., smoking, protein levels) are considered in place of individual DNAm CpG sites that are typically used to derive first-generation clocks. While this incorporation of broader biological information may enhance the utility for clinical outcome prediction, it also complicates interpretability compared to first-generation clocks, which may remain more suitable for investigating the mechanisms underlying cellular ageing.

A key question in evaluating the clinical utility of epigenetic clock biomarkers of aging is their capacity to predict the future incidence of aging-related disease. However, while there are now a large number of studies reporting clock associations with different disease outcomes in a range of datasets [9], [10] no large-scale systematic comparison of many clocks predicting many diseases has been conducted.

Here, we provide an unbiased assessment of 14 leading clocks (**Table 1**) in relation to 10-year onset of a comprehensive set of 174 disease outcomes within one of the world’s single-largest DNAm datasets, the Generation Scotland cohort (n=18,859) [11]. There was a minimum of 30 incident cases per disease (median=108, max=1,527 cases for hypertension, see **Table S1**).

**Table 1.** Overview of the 14 epigenetic clocks considered as disease risk predictors.

| Clock | Generation | Outcome | Tissue | N CpGs in Clock |
| --- | --- | --- | --- | --- |
| Hannum (2013) [4] | First | Age (years) | Blood | 71 CpGs |
| Horvath (2013) [5] | First | Age (years) | Multi- tissue | 353 CpGs |
| Lin (2016) [18] | First | Age (years) | Blood | 100 CpGs |
| Zhang_10 (2017) [19] | Second | Mortality | Blood | 514 CpGs |
| PhenoAge (2018) [6] | Second | Phenotypic Age | Blood | 513 CpGs |
| Horvath Skin & Blood (2018) [20] | First | Age (years) | Skin and Blood | 391 CpGs |
| GrimAge v1 (2019) [7] | Second | Mortality | Blood | 1,030 CpGs |
| DNAmTL (2019) [21] | NA | Telomere Length | Blood, Adipose | 146 CpGs |
| Dunedin PoAm38 (2020) [22] | Third | Pace of Ageing | Blood | 46 CpGs |
| Dunedin PACE (2022) [8] | Third | Pace of Ageing | Blood | 173 CpGs |
| GrimAge v2 (2022) [23] | Second | Mortality | Blood | 1,030 CpGs |
| YingCausAge (2024) [24] | First | Age (years) | Blood | 586 Causal CpGs |
| YingDamAge (2024) [24] | First | Age (years) | Blood | 1,090 Damaging CpGs |
| YingAdaptAge (2024) [24] | First | Age (years) | Blood | 1,000 Protective CpGs |

Cox proportional hazards regression was run for each clock-disease pairing, adjusting for age, sex, body mass index, smoking, alcohol consumption, education, and socioeconomic deprivation (**Table S2**). Age, estimated cell proportions and relatedness were regressed out of each clock prior to the Cox regression analyses. This covariate setup might directly capture information that is included in the clocks e.g., a DNAm proxy for smoking is included in the construction of GrimAge, a predictor of mortality [7]. However, these covariates are easy to assess and commonly collected in clinical settings. Thus, in terms of disease prediction, it is valuable to note what added information epigenetic clocks might contribute. A total of 176 Bonferroni significant associations were found for 13 of the clocks across 57 diseases (P < 2.9 x10^−4^). In addition to the Cox regressions, logistic regression 10-year classification was conducted using the same covariates (null model). The differences in AUC between these models and full models (adding in an epigenetic clock) were then calculated.

An R ShinyApp displays the results from both the Cox and logistic regression analyses (https://shiny.igc.ed.ac.uk/Epigenetic_Clock_and_Disease_Association/). Users can visualise findings for specific or multiple clocks and disease outcomes or diseases within a grouping e.g, psychiatric or cardiovascular traits.

There were 9 Bonferroni significant disease associations for the first-generation clocks, representing ~5% of all significant findings. Across all 174 diseases that were considered, and using GrimAge v1 as the reference category, the average log hazards (**Figure 1, Table S3 and S4**) of the first-generation clocks were around 50% smaller in magnitude (P≤2.9×10^−10^). Smaller effect sizes were also noted for PhenoAge (P=6.3×10^−3^) and DNAm telomere length (P<2×10^−16^). There were no significant differences with the remaining second generation or third generation clocks (GrimAge v2, DunedinPoAm and DunedinPACE; P>0.05) apart from Zhang10 (P=0.042).

**Figure 1.**
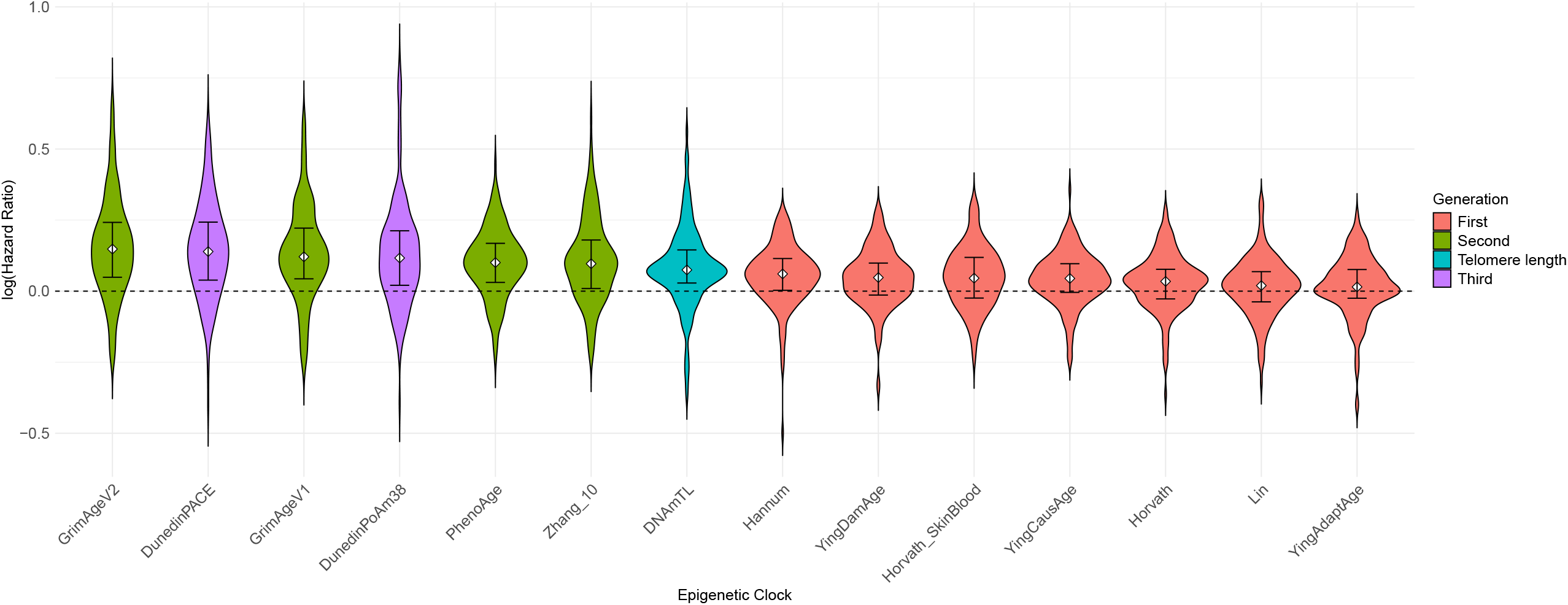
Distribution of log hazard ratios for each epigenetic clock across all 174 incident disease outcomes. Clocks are presented in descending order of the median effect size. Median and interquartile range presented within the violins. First generation clocks are highlighted in pink, second generation in green, third generation clocks in purple and telomere length (where log HR effect sizes have been multiplied by −1 for visual display purposes) in turquoise.

To help contextualise the magnitude and significance of the clock-disease associations, we first ran survival models for 10-year all-cause mortality (n_cases_ = 842; **Table S2**) – one of the best-established health outcomes to be related to epigenetic age acceleration [12]. The largest and most significant association was with GrimAge v2 (Hazard Ratio (HR) per SD of age acceleration = 1.54, 95% CI [1.46, 1.62], P = 7.1×10^−62^). All clocks had significant associations at P<0.05 apart from Horvath’s skin and blood clock, Ying’s AdaptAge and Lin’s age predictor. Using logistic regression, an AUC of 0.851 was obtained for the model with covariates only. This increased by up to 0.014 (1.4%) upon the addition of GrimAge v2 (AUC=0.865) as a covariate.

There were 27 unique disease outcomes where the clock-disease hazard ratio was Bonferroni-significant and exceeded the magnitude of the corresponding clock-mortality association (**Table S5**). A diverse set of diseases were highlighted, however there was a strong focus on respiratory/smoking-related and liver-related outcomes, including primary lung cancer (HR_GrimAgev1_ per SD = 1.56 [1.42, 1.72], P = 5.3×10^−19^) and cirrhosis (HR_GrimAgev2_ = 1.86 [1.57, 2.21], P = 8.9×10^−13^). Also of note, were associations with diabetes (HR_DunedinPACE_ = 1.44 [1.33, 1.57], P = 9.6×10^−19^), Crohn’s disease (HR_PhenoAge_ = 1.39 [1.19, 1.64], P = 4.7×10^−5^) and delirium (HR_Zhang10_ = 1.44 [1.23, 1.68], P = 6.7×10^−6^).

Logistic regression models were then run for all clock-disease analysis using a null model (covariates only) and a full model where a clock was added as a predictor of interest. After filtering to the 176 clock-disease associations that were Bonferroni significant in the Cox models, there were 35 instances where the AUC from the full model exceeded 0.8 (indicative of being clinically meaningful) and the AUC improvement between the null and full model was greater than 0.01 (**Figure 2** and **Table S6**).

**Figure 2.**
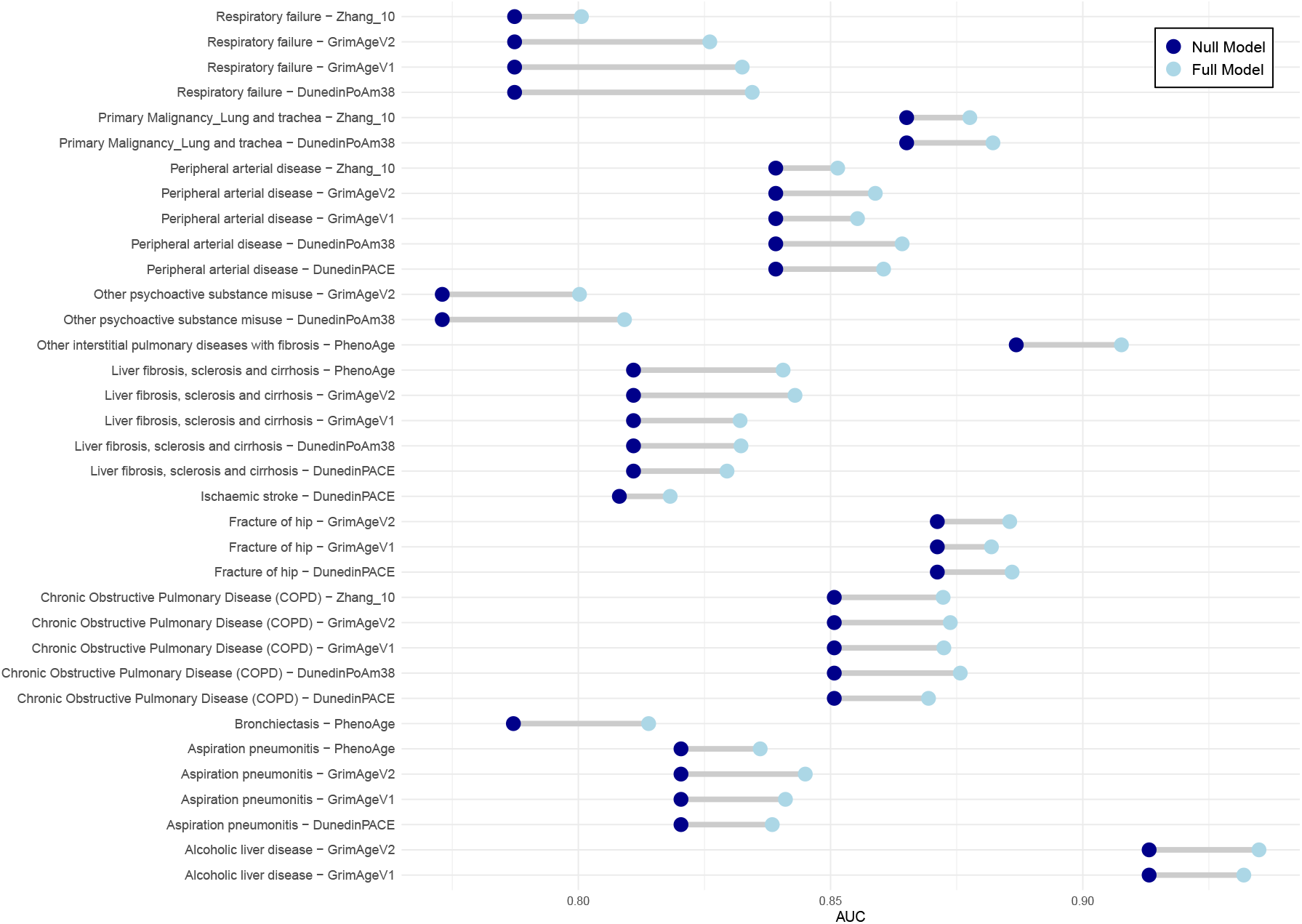
AUC increments for disease – clock associations where the hazard ratio from the Cox regression was Bonferroni significant, the AUC of the full model was >0.8 and the improvement to the AUC upon adding the clock to the null model was >0.01. Null model covariates include: age, sex, education, alcohol, smoking, BMI and deprivation. The full models also included the relevant epigenetic clock.

Again, several clocks were linked to respiratory/smoking and alcohol-related disease outcomes, including COPD, primary oesophageal and lung cancer, respiratory failure and cirrhosis. In addition, PhenoAge and DunedinPACE improved classification of Parkinson’s disease and ischaemic stroke, respectively. Hannum’s clock and Horvath’s skin and blood clock improved the classification of primary uterine and pancreatic cancer, respectively.

Our findings clearly indicate that second- and third-generation epigenetic clocks should be prioritised for disease association studies. These clocks showed particularly strong links to respiratory and liver-related disease outcomes, including primary lung and oesophageal cancers and cirrhosis. Furthermore, the findings are present after adjusting for key risk factors such as deprivation, self-reported smoking behaviour and alcohol consumption as covariates.

The 162 Bonferroni significant second- and third-generation clock disease associations were evenly spread across the clocks, with a maximum of 37 associations from GrimAge v2 i.e., there is no clear “best” clock for pan-disease analyses. The construction of both v1 and v2 of GrimAge includes a DNAm surrogate for smoking, which may help to drive downstream associations with respiratory diseases. Similarly, the inclusion of longitudinal changes in BMI, waist-hip ratio and HbA1c in DunedinPACE may contribute to its predictive utility for diabetes.

Our study contains certain limitations. For example, we related DNAm profiling from whole-blood samples against multi-tissue diseases and non-blood tissue specific diseases in a Scottish-based cohort. Our data analysis did not exhaustively adjust for covariates that might track disease risk and multi-morbidity, such as family history of disease, genetic risk scores (polygenic scores), medication usage, social interactions and blood pressure. However, bespoke covariate setups are not feasible when studying such a broad range of disease outcomes. Smoking status and alcohol consumption data were self-reported and may therefore carry biases. Future studies could consider the addition of epigenetic proxies for these measures as covariates. It is also crucial to highlight that epigenetic clocks do not provide insights into the mechanisms of action of diseases. Lastly, despite the longitudinal data utilized in this study to compute incident disease outcomes, serial biosamples were not available.

Replication of our findings using longitudinal samples across diverse cohorts is needed. The time from blood draw to disease onset varied in this study. Investigations of specific time windows e.g., in increments of 5 years, could yield different strengths of associations. Future research should also consider the training and testing of disease-specific clocks. For example, we have previously shown that a DNAm predictor for incident Type 2 Diabetes augments models containing traditional risk factors [13]. Finally, our disease phenotyping approach, whilst consensus-based, might be too broad in some instances e.g., the all-cause dementia phenotype captures both Alzheimer’s disease and vascular dementia, as well as other, less common dementia sub-types.

Here, we performed a comprehensive and unbiased comparison of leading epigenetic clocks as putative biomarkers for 174 incident disease outcomes. We identified 176 disease associations across 13 clocks with 57 unique diseases. By anchoring our findings to well-established associations with all-cause mortality, we highlighted multiple novel associations with large effect sizes. We also focus on outcomes where classification gains may lead to clinical impact. These results form a starting point for the targeted selection of epigenetic clocks for consideration in clinical risk prediction models.

## Methods

### Generation Scotland Cohort

Generation Scotland (GS) is a family-based research study comprised of 24,084 volunteers [14]. Volunteers joined the cohort between 2006 and 2011 with the majority of recruitment taking place via invitation from the individuals’ general practitioners (GPs). Initially, individuals aged 35-65 years were recruited from across five geographic areas (Glasgow, Aberdeen, Dundee, Ayr and Arran). They were then encouraged to recruit family members living in Scotland to join the study. This resulted in a baseline cohort aged between 17 and 99 years. All individuals were invited to complete questionnaires and to attend a baseline clinic visit for further questionnaires and testing, resulting in a detailed collection of lifestyle, clinical, health, cognitive and sociodemographic data. The majority also agreed to provide biosamples, including blood, from which DNA has been extracted and profiled for genetic and epigenetic data. Most individuals also provided consent for researchers to access their medical records via data linkage to primary and secondary care codes.

### Ethics and Consent

All components of Generation Scotland received ethical approval from the NHS Tayside Committee on Medical Research Ethics (REC Reference Number: 05/S1401/89). All participants provided broad and enduring written informed consent for biomedical research. Generation Scotland has also been granted Research Tissue Bank status by the East of Scotland Research Ethics Service (REC Reference Number: 15/0040/ES), providing generic ethical approval for a wide range of uses within medical research. This study was performed in accordance with the Helsinki declaration.

### DNA Methylation Profiling

Methylation data have been quantified for 18,413 GS volunteers at a total of 752,722 genomic (CpG) sites after quality control via the Illumina EPIC850k array [11]. Thereafter, epigenetic age and age acceleration estimates were derived via the Biolearn platform. Biolearn is an open-source python library that enables easy and versatile analyses of biomarkers of aging data. It provides tools to easily load data from publicly available sources and contains reference implementations for common aging clocks such as the Horvath clock, DunedinPACE, and many others that can easily be run in only a few lines of code [15].

Age acceleration for each clock/predictor was calculated as the residual from a linear mixed model where the clock/predictor was regressed on age and estimated cell proportions (derived from the Biolearn platform). To avoid collinearity (as the cell proportions sum to one), basophil proportion was not included as a covariate. A pedigree kinship matrix was included as a random effect to control for the family structure present in the cohort. The regression models were run using the lmekin function from the *coxme* R package [16].

### Disease Coding

Using a list of consensus definitions based on primary and secondary care codes [17], 308 disease outcomes were derived from the electronic health records data in Generation Scotland from January 1980 to April 2022. Despite having volunteer consent to access primary care data, these were only accessible for ~40% of the cohort due to consent constraints with individual GP surgeries (the data holders). These data were subsequently filtered to only consider the first diagnosis for each disease, which could be made in either a primary or secondary care setting. Those with a diagnosis prior to joining the study were excluded from downstream analyses for that specific disease outcome, in order to ensure investigation of incident as opposed to prevalent disease outcomes. Not all diseases were observed in the cohort. The latest date of linkage with primary and secondary healthy records was April 2022. Filtering to diseases with at least 30 cases over the first 10-years of follow-up resulted in 174 outcomes for the main analyses. A list of the 174 diseases observed and the number of incident events (events occurring after the blood draw) along with mean age at diagnosis are presented in **Table S1**.

### Mortality Records

Mortality records were obtained via Community Health Index linkage to the National Records of Scotland. These data formed part of our censoring criteria where individuals either survived and remained unaffected by a disease or died during the follow up period to April 2022.

### Covariates

Demographic, lifestyle and health variables were considered as covariates in the regression models with each clock and disease outcome. These included: age, sex, body mass index (BMI, calculated as weight in kg divided by squared height in metres), smoking pack years (calculated by self-reported packs per day multiplied by number of years as a smoker), alcohol units consumed over the last week, years of education (an ordinal variable with 11 categories: (1) 0 years, (2) 1–4 years, (3) 5–9 years, (4) 10–11 years, (5) 12–13 years, (6) 14–15 years, (7) 16–17 years, (8) 18–19 years, (9) 20–21 years, (10) 22–23 years, (11) 24 or more years), and an area-based rank of socioeconomic deprivation (Scottish Index of Multiple Deprivation, SIMD). SIMD measures relative deprivation across 6,976 data zones, calculated across seven domains: income, employment, education, health, access to services, crime and housing.

K-nearest neighbours’ imputation – using k=10 and a maximum missingness per person of 4/7 of variables – was used to generate complete data for covariates. There was complete data on age and sex with a range of missingness for the other variables (SIMD: n = 1,152, BMI: n = 120, Education: n = 1,012, Pack years: n = 395, Alcohol: n = 1,734). Natural log transformations were applied to alcohol and pack years (adding a constant of 1 to all values to account for non-drinkers/smokers) and BMI. All covariates were then standardised to mean zero and unit variance prior to imputation.

### Association between age acceleration and disease outcomes

Cox proportional hazards regression models, adjusting for the aforementioned covariates, were used to relate age acceleration for each epigenetic clock to 10-year onset for the list of 174 incident disease outcomes. The analyses were subset to the most frequently affected by sex where the proportion of male or female cases was >90% for any given disease. A total of 174 models were run for each of the 14 clocks, giving 2,436 models in total. P-value filtering was conducted per individual clock-regression model, resulting in a Bonferroni threshold of P < 0.05/174 = 2.9 × 10^−4^. A further set of analyses were run where time to all-cause mortality was the outcome.

The proportional hazards assumption was tested by examining the Schoenfeld residuals. A test of the residuals for both the overall model and the predictor variable of interest (age acceleration) was performed using the cox.zph function in the survival package.

A formal comparison of the log Hazard Ratios for the clocks across all 174 disease outcomes was then conducted using a linear mixed effects model. The logHR was the outcome while the clock was the predictor (GrimAge v1 was the reference level) and disease was included as a random effect on the intercept.

In addition to the Cox models, logistic regression was used to generate area under the curve (AUC) estimates for 10-year disease classification. Null models contained the aforementioned covariates, while a full model also included an age acceleration measure.

## Code and Data Availability

Fully annotated copies of the analysis scripts are available at: https://github.com/marioni-group/Epigenetic_Clocks_and_Disease_GS.

The source data that were analysed during the current study are not publicly available due to them containing information that could compromise participant consent and confidentiality. Data can be obtained from the data owners. Instructions for accessing Generation Scotland data can be found here: https://www.ed.ac.uk/generation-scotland/for-researchers/access; the ‘GS Access Request Form’ can be downloaded from this site. Completed request forms must be sent to to be approved by the Generation Scotland Access Committee.

## Ethics approval and consent to participate

All components of Generation Scotland received ethical approval from the NHS Tayside Committee on Medical Research Ethics (REC Reference Number: 05/S1401/89). GS has also been granted Research Tissue Bank status by the East of Scotland Research Ethics Service (REC Reference Number: 20-ES-0021), providing generic ethical approval for a wide range of uses within medical research.

All participants provided written informed consent. The study was performed in accordance with the Helsinki declaration.

## Acknowledgements

This research was funded in whole, or in part, by the Wellcome Trust (104036/Z/14/Z and 221890/Z/20/Z). For the purpose of open access, the author has applied a CC BY public copyright license to any Author Accepted Manuscript version arising from this submission. Generation Scotland received core support from the Chief Scientist Office of the Scottish Government Health Directorates (CZD/16/6) and the Scottish Funding Council (HR03006). DNA methylation profiling of the Generation Scotland samples was carried out by the Genetics Core Laboratory at the Edinburgh Clinical Research Facility, Edinburgh, Scotland, and was funded by the Medical Research Council UK and Wellcome (Wellcome Trust Strategic Award STratifying Resilience and Depression Longitudinally (STRADL; Reference 104036/Z/14/Z). DNA methylation data for Generation Scotland was also funded by a 2018 NARSAD Young Investigator Grant from the Brain & Behavior Research Foundation (Ref: 27404; awardee: Dr David M Howard) and by a John, Margaret, Alfred and Stewart Sim Fellowship from the Royal College of Physicians of Edinburgh (Awardee: Dr Heather C Whalley).

D.W.B a fellow of the CIFAR CBD Network and is supported in part by R01AG073402 and R01AG087158. T.C and C.M are funded by the Mayo Clinic Robert and Arlene Kogod Center on Aging.

## List of Supplementary Tables

**Table S1**. Descriptive statistics for the 174 disease outcomes

**Table S2**. Cox regression and logistic regression results for 14 clocks and 174 incident disease outcomes plus all-cause mortality.

HR = Hazard Ratio, LCI = lower 95% confidence interval, UCI = upper 95% confidence interval, P = p-value, Schoenfeld local p = p-value for the Shoenfeld residual test for the predictor (clock) of interest, Schoenfeld global p = p-value for the Shoenfeld residual test for all predictors in the model, auc null = area under the curve for the logistic regression model with covariates only, auc full = area under the curve for the logistics regression model with covariates and the clock of interest, auc diff = difference between the auc_null and auc_full

**Table S3**. Mean and standard deviation of the log hazard ratios for each clock, averaged across all 174 disease associations.

**Table S4**. Linear mixed effects model output contrasting the mean log hazard ratio for each clock against GrimAge v1 (reference category).

**Table S5**. Subset of Table S2 showing Bonferroni significant associations where the log hazard ratio is greater for the respective disease-clock association compared to the mortality-clock association.

**Table S6**. Subset of Table S2 showing Bonferroni significant associations where the AUC of the fully-adjusted model exceeded 0.8 and the difference in AUC with the basic-adjusted model is greater than 0.01 (1%).

